# A Statistical and Dynamical Model for Forecasting COVID-19 Deaths based on a Hybrid Asymmetric Gaussian and SEIR Construct

**DOI:** 10.1101/2020.06.21.20136937

**Authors:** Jack A. Syage

**Affiliations:** ImmunogenX, 1600 Dove Street, Suite 330, Newport Beach, CA 92660

## Abstract

**Background:** The limitations of forecasting (real-time statistical) and predictive (dynamic epidemiological) models have become apparent as COVID-19 has progressed from a rapid exponential ascent to a slower decent, which is dependent on unknowable parameters such as extent of social distancing and easing. We present a means to optimize a forecasting model by functionalizing our previously reported Asymmetric Gaussian model with SEIR-like parameters. Conversely, SEIR models can be adapted to better incorporate real-time data.

**Methods:** Our previously reported asymmetric Gaussian model was shown to greatly improve on forecasting accuracy relative to use of symmetric functions, such as Gaussian and error functions for death rates and cumulative deaths, respectively. However, the reported asymmetric Gaussian implementation, which fitted well to the ascent and much of the recovery side of the real death rate data, was not agile enough to respond to changing social behavior that is resulting in persistence of infections and deaths in the later stage of recovery. We have introduced a time-dependent σ(t) parameter to account for transmission rate variability due to the effects of behavioral changes such as social distancing and subsequent social easing. The σ(t) parameter is analogous to the basic reproduction number R_0_ (infection factor) that is evidently not a constant during the progression of COVID-19 for a particular population. The popularly used SEIR model and its many variants are also incorporating a time dependent R_0_(t) to better describe the effects of social distancing and social easing to improve predictive capability when extrapolating from real-time data.

**Results:** Comparisons are given for the previously reported Asymmetric Gaussian model and to the revised, what we call, SEIR Gaussian model. We also have developed an analogous model based on R_0_(t) that we call SEIR Statistical model to show the correspondence that can be attained. It is shown that these two models can replicate each other and therefore provide similar forecasts based on fitting to the same real-time data. We show the results for reported U.S. death rates up to June 12, 2020 at which time the cumulative death count was 113,820. The forecasted cumulative deaths for these two models and compared to the University of Washington (UW) IHME model are 140,440, 139,272, and 149,690 (for 8/4/20) and 147,819, 148, 912, and 201,129 (for 10/1/20), respectively. We also show how the SEIR asymmetric Gaussian model can also account for various scenarios of social distancing, social easing, and even rebound outbreaks where the death and case rates begin climbing again.

**Conclusions:** Forecasting models, based on real-time data, are essential for guiding policy and human behavior to minimize the deadly impact of COVID-19 while balancing the need to socialize and energize the economy. It is becoming clear that changing social behavior from isolation to easing requires models that can adapt to the changing transmission rate in order to more accurately forecast death and case rates. We believe our asymmetric Gaussian approach has advantages over modified SEIR models in offering simpler governing equations that are dependent on fewer variables.

## INTRODUCTION

The progression of COVID-19 in almost all populations around the world is dependent on social behavior such as distancing followed by easing. This presents a challenge to models that attempt to forecast important properties such as case and death rates. There are two general categories of modeling:

- The traditional approach is epidemiological in nature and seeks to understand known behavior, which for an epidemic, would be after the fact. Conventional epidemiological models include SIR, SEIR and SEIRS (susceptible, exposed, infectious, recovered, susceptible) models^1,2^ and are characterized by mathematical complexity and dependence on many variables that can be difficult to define.^3, 4^ They are also not well suited to real-time forecasting as new epidemics have their own distinct properties and further it is difficult for them to consider implementation of social restrictions or changes in social behavior due to government easing and/or public rebellion due to economic and social pressures.
- For the purposes of forecasting, an alternative to epidemiological parameters and coupled kinetic equations is to use shape functions that are fitted to real-time data for cases and deaths (and can incorporate other measurables) in order to extrapolate to future dates. Several real-time statistical models have emerged that specifically address the unique properties and behavior of COVID-19, knowledge that is only learned by following daily trends.^5,6,7,8^

The limitations of predictive (dynamic epidemiological) and forecasting (real-time statistical) models have become very apparent as COVID-19 has progressed from a rapid exponential ascent to a slower decent that is dependent on unpredictable behavior such as extent of social isolation and easing. We present a means to optimize a forecasting model by functionalizing our previously reported Asymmetric Gaussian model^9,10^ with SEIR-like parameters. Conversely, SEIR models can be adapted to better incorporate real-time data, but these hybrid models still appear to be dependent on many difficult-to-define variables and are very complex.^11,12,13,14,15^

In this paper, we extend the applicability of our asymmetric Gaussian model^9,10^ by instilling it with SEIR-like parameters, such as transmission rate R_0_, and more importantly allowing this value to vary depending on the extent and timing of social distancing and easing in a given population. This variability is incorporated into a time-dependent sigma value σ(t) for our asymmetric Gaussian model. Our model is strongly dependent on real-time death-rate data as we believe that these data are the most reliably reported. The use of case data we consider much less reliable since it is convoluted with the extent of testing for a given population and given that this changes with time, the shape of the case-rate plot is not representative of the true shape, which is imperative for achieving reasonable forecasting. A widely followed model for COVID-19 is from the Institute for Health Metrics and Evaluation (IHME) at the University of Washington (UW).^5^ As best as we can determine the IHME model uses the integrated total death curve (S-curve) to determine where on the time progression a population lies in order to integrate to total deaths. Alternatively, we use the death-rate curve (1^st^ derivative of the total death curve) as it is more sensitive to detection of the inflection point for peak death rate.^9,10^ Further we recognized early that a symmetric Gaussian was an ill fit particularly on the descending recovery side. The IHME model fitted the cumulative death curve to an error function (integral to a symmetric Gaussian) and consistently underestimated future deaths until adopting a more SEIR like approach recently.^16^

We believe our asymmetric Gaussian approach has advantages over modified SEIR models in offering simpler governing equations that are dependent on fewer variables.

## METHOD

The basic premise of our model has been reported before.^9,10^ Briefly, the general algorithm uses death-rate data and an asymmetric Gaussian rise and fall curve to forecast total deaths by estimating where on the model curve the actual death-rate curve lies. From that time point one can integrate remaining time to forecast a final total death count. By also using measured values for the time from infection to recovery or death and a mortality factor, the prevalence (active cases) and incidence (new cases) totals and rate curves can be constructed. It is also possible by setting a downward threshold on prevalence that an estimate of a minimum date to relax social restrictions may be considered. Other functions besides a Gaussian could work (and we discussed some of them before)^9^ but they don’t have mathematical simplicity and the COVID-19 data currently doesn’t provide a compelling reason to consider more complicated functions. On March 14, 2020 we launched a website blog to report our developing model for COVID-19.^10^

Our previously reported asymmetric Gaussian model greatly improved forecasting on the recovery side of the death rate curve, but was not agile enough to respond to changing social behavior that is that is resulting in persistence of infections and deaths in the later stage of recovery. As alluded to in our earlier paper we have now implemented a functionalized sigma value σ(t) that is descriptive of the changing infection or reproduction rate generally represented by R_0_, which has become evident is not a constant during the progression of the disease for a particular population.

The operating equations for the asymmetric Gaussian model are as follows:

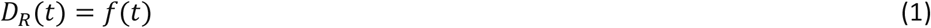

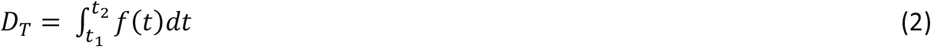

Where *D*_*R*_ and *D*_*T*_ are death rates (e.g., daily) and total deaths, respectively. We define *f(t)* as an asymmetric Gaussian function by way of a time-dependent *σ(t)* parameter that represents changing transmission rates due to timing of social restrictions and subsequent easing.

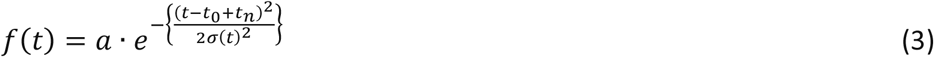

where

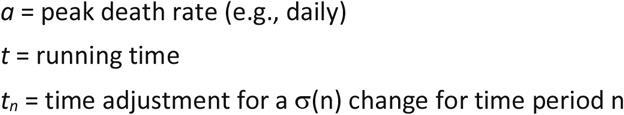

The integration of f(t) and the determination of other time dependences such as cases and incidences have been presented before^9^ and are routinely calculated in simple programs such as Excel. Rather than choose a function for σ(t) we instead define discrete values σ(n) for different time periods n along the COVID-19 death rate progression reflecting changing social behavior such as initial unrestricted exponential growth followed by social distancing and then by social easing. One complication of this approach is that an abrupt change in σ will cause an abrupt change in f(t). We handle this transition by proportionately changing the overall time in Eq. (3) through the term t_n_ such that

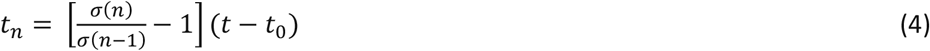

Alternatively, we could define a σ(t) that continuously changes; however, defining such a function would be unwieldy. One could also define discrete values for different time periods and create a smooth curve with an appropriate running average. However, we prefer to use the transition factor of Eq. (4) to minimize any discontinuities at the transition points.

We now show how a SEIR model can be modified to also incorporate discrete values for different times for R_0_. The intent here is to show that a statistical model such as the one reported here can replicate an epidemiological model such as SEIR. The governing equations for standard SEIR are well known and published and won’t be repeated here. If we ignore birth and death rates then β = R_0_/r where β is the contact rate, r is the recovery (removal) time and R_0_ is the basic reproduction number. We consider R_0_ to be time dependent in order to develop an analog to our time dependent σ(t) for the asymmetric Gaussian model. In this way we show that the asymmetric Gaussian model with functionalized sigma can replicate the variable R_0_ SEIR model.

## RESULTS

Table I shows the input parameters for the previously reported Asymmetric Gaussian model, the current functionalized sigma model, which for lack of a better name we refer to as SEIR Gaussian, and a SEIR model with time varying R_0_ that can fit to real-time data, which again for lack of a better name we refer to as SEIR Statistical. The values in parentheses represent the three time periods approximately representing the epidemic before social distancing (time period 1), social distancing (time period 2), and social easing (time period 3), and τ(n,n+1) represents the time for transition between these time periods (n = 1, 2, 3). These parameters were obtained as best fit values from visual inspection. We fixed values of r and m (d is not required for death plots, but is used for calculating prevalence and incidence). We also tried to make the τ(n,n+1) parameter as close between the SEIR Gaussian and SEIR Statistical model to attempt to show correspondence. The Gaussian σ and the SEIR R_0_ parameters are related, but there is no clear functional relationship between the two, which is not unexpected since they plug into very different governing equations. So, we treat them as empirical parameters to fit to the real-time data.

**Table I.** Input parameters for statistical model comparison. (σ and τ values are in units of days.)

|  |  | Asymmetric<br>Gaussian | SEIR<br>Gaussian | SEIR<br>Statistical |
| --- | --- | --- | --- | --- |
| Recovery (days) | $r =$ | 24.7 | 24.7 | 24.7 |
| Death (days) | $d =$ | 17.8 | 17.8 | |
| Incubation (days) |  |  |  | 6 |
| Mortality | $m =$ | 1.0% | 1.0% | 1.0% |
| Transmission | $R_0(1)$ | | | 6.0 |
| | $R_0(2)$ | | | 0.4 |
| | $R_0(3)$ | | | 0.55 |
| | $\sigma(1)$ | 10 | 10 | |
| | $\sigma(2)$ | 32 | 27 | |
| | $\sigma(3)$ | | 84 | |
| | $\tau(1,2)$ | 0 | 0 | -8 |
| | $\tau(2,3)$ | | 32 | 32 |

The best fits of these three models to reported U.S. death rate D_R_ and cumulative death D_T_ statistics are plotted in Figure 1. The key observations are:

**Figure 1.**
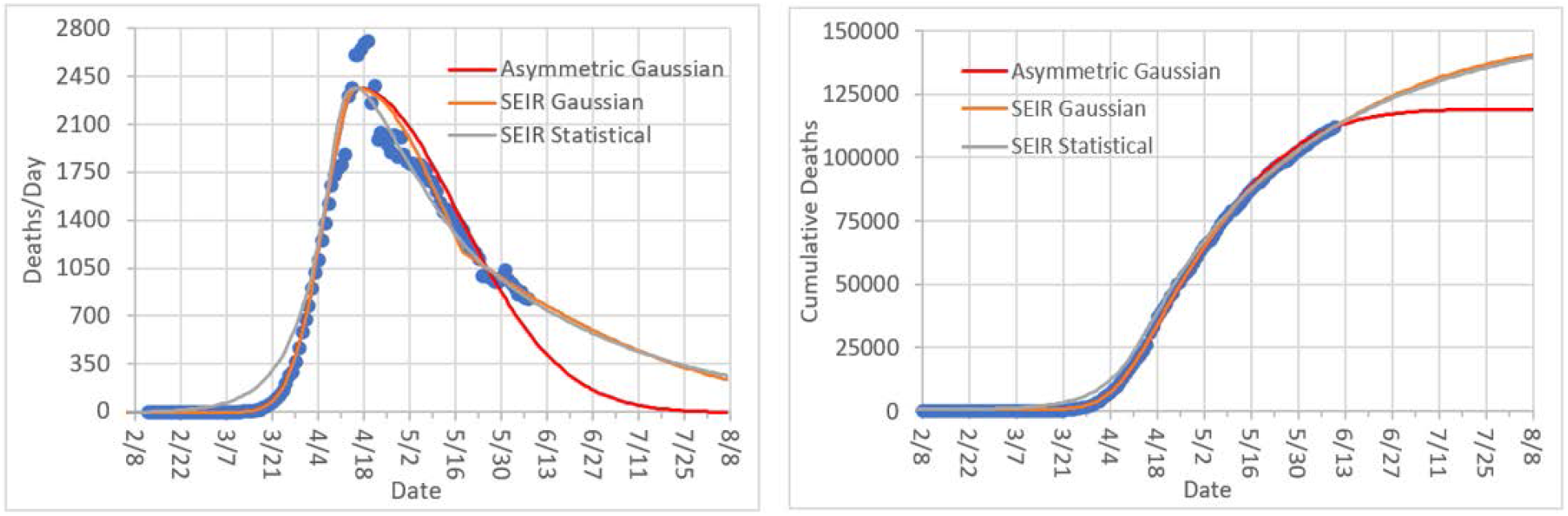
Data for U.S. death rate up to 6/12/20 and corresponding curve fits for the three statistical models considered here. The cumulative death count on this date was 113,820.^17^

- The Asymmetric Gaussian (red curve) does well fitting to the rise and about halfway down the fall. However, it does not forecast the persistence in the decline of the death rate. This is also seen in the cumulative death data.
- The SEIR Gaussian and SEIR Statistical models forecast nearly exactly the same for the parameters in Table I. The former model may be a little better at the onset of the epidemic as can be seen by a premature rise by the latter model, but this is inconsequential when integrating over the entire death rate curve.

We now summarize the forecast of total deaths at various future dates in Table II. The intent here is to show that the simpler SEIR Gaussian model can replicate the forecasting of the more complicated SEIR Statistical model as evident in Table II and Figure 1. The UW IHME model, which has consistently under forecasted in the past, underwent a few weeks ago a change in their algorithm and now shows rampant growth in certain populations (such as CA) that results in much higher forecasted values for all of the U.S.

**Table II.** Forecasted total deaths for future dates by the three reported models and by the UW IHME model.

|  | Asymmetric<br>Gaussian | SEIR<br>Gaussian | SEIR<br>Statistical | UW IHME |
| --- | --- | --- | --- | --- |
| 8/4/2020 | 120,642 | 140,440 | 139,272 | 149,690 |
| 9/5/2020 | 120,699 | 145,852 | 145,749 | 172,138 |
| 10/1/2020 | 120,700 | 147,819 | 148,912 | 201,129 |

The SEIR (asymmetric) Gaussian model can also account for various scenarios of social distancing, social easing, and even rebound outbreaks where the death and case rates begin climbing again. Figure 2 shows various scenarios for the parameters σ(n) and τ(n,n+1). The yellow curve (1^st^ legend row) represents the fitted curve to the U.S. data in Figure 1. The blue curve (2^nd^ row) shows a slower declining curve when changing σ(3) from 84 to 200 reflecting greater social easing. The purple curve (3^rd^ row) shows a rebound outbreak in the social easing time period where now σ(3) = −40. For this curve we also show the effect of increasing σ(2) in the social distancing time period from 27 to 32 representing less effective social distancing. Finally, the gray curve (4^th^ row) shows the effect of a delay in social easing by 10 days, i.e., τ(2,3) increases from 32 to 42 days. This provides a capability to inform government agencies the consequences of deciding when to implement social easing. A shortcoming of this model is evident by the abrupt changes in the curve at each time period transition point. However, this can be remedied by applying an appropriate amount of smoothing (i.e., running average). Another shortcoming in its present form is the rebound outbreak modeling reaches the same peak as the first outbreak. These are treatable by either applying an overall intensity factor I(t) in place of the *a* term in Eq. (3) or changing the calculation for t_n_ in Eq. (4), however this adds another variable to the fitting parameters.

**Figure 2.**
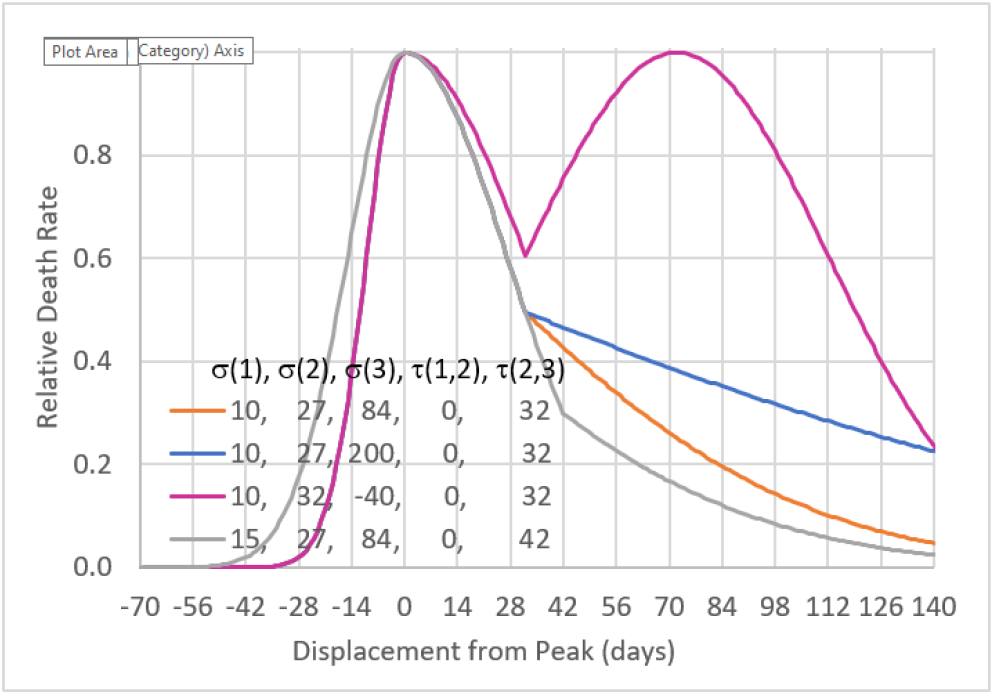
The SEIR Gaussian model for varying σ(n) and τ(n,n+1).The first (yellow) curve are for the parameters that fit the U.S. date in Figure 1.

The main conclusion from the above results is that the SEIR Gaussian model can offer comparable forecasting power as the SEIR Statistical model. However, our SEIR Gaussian model uses a single function and only needs the variables σ(1), σ(2), σ(3) and τ(2,3). The value τ(1,2) can be fixed at 0 and the only other variable used, d, is for calculating and forecasting of case prevalence and incidence. The SEIR Statistical Model requires solving five coupled differential equations and requires the variables r, R_0_(1), R_0_(2), R_0_(3), τ(1,2) and τ(2,3).

## DISCUSSION

The above results describe a forecasting model that is simple in principle, but with capability to address more subtle dynamics of the on-going COVID-19 pandemic. We now discuss some of these present and future refinements. We first note that fitting the R_0_(t) version of the SEIR model to the U.S. death rate data leads to forced values of R_0_ that seem unrealistic, e.g., higher and lower than expected values, respectively for R_0_(1) and R_0_(3). Rather than try to explain this we treat them instead as empirical fitting parameters as that is how we treat the analogous σ(n) parameters for the SEIR Gaussian model.

Although it might seem like a misnomer to apply that adage SEIR to our latest asymmetric Gaussian model, in fact the time dependence of each of these compartments, namely S, E, I, R, and D, are calculable using this model, just not with kinetic equations. The SEIR Gaussian model is arguably better-suited for epidemics such as COVID-19 where herd immunity is not happening to a great extent than SEIR models, which seem more designed for describing situations where a large fraction of the susceptible population become infectious. Ultimately what is important is to derive a shape function that fits to the available real-time data and extrapolates as accurately as possible to values at future dates.

## CONCLUSION

Forecasting models, based on real-time data, are essential for guiding policy and human behavior to minimize the deadly impact of COVID-19 while balancing the need to socialize and energize the economy. It is becoming clear that changing social behavior from isolation to easing requires models that can adapt to the changing transmission rate in order to more accurately forecast death and case rates. We believe our asymmetric Gaussian approach has advantages over modified SEIR models in offering simpler governing equations that are dependent on fewer variables without any evident accuracy and precision penalties.

## Data Availability

All data are available upon request. Some of this work has been reported on:
syage-covid19-assessment.com

https://syage-covid19-assessment.com

